# Daily Lactobacillus Probiotic versus Placebo in COVID-19-Exposed Household Contacts (PROTECT-EHC): A Randomized Clinical Trial

**DOI:** 10.1101/2022.01.04.21268275

**Authors:** Paul E. Wischmeyer, Helen Tang, Yi Ren, Lauren Bohannon, Zeni E. Ramirez, Tessa M. Andermann, Julia A. Messina, Julia A. Sung, David Jensen, Sin-Ho Jung, Alexandra Artica, Anne Britt, Amy Bush, Ernaya Johnson, Meagan V. Lew, Hilary M. Miller, Claudia E. Pamanes, Alessandro Racioppi, Aaron T. Zhao, Neeraj K. Surana, Anthony D. Sung

## Abstract

**Importance:** The COVID-19 pandemic continues to pose unprecedented challenges to worldwide health. While vaccines are effective, supplemental strategies to mitigate the spread and severity of COVID-19 are urgently needed. Emerging evidence suggests susceptibility to infections, including respiratory tract infections, may be reduced by probiotic interventions; therefore, probiotics may be a low-risk, widely implementable modality to mitigate risk of COVID-19 disease, particularly in areas with low vaccine availability and/or uptake.

**Objective:** To determine whether daily probiotic *Lactobacillus rhamnosus* GG (LGG) is effective in preventing development of symptoms of illness within 28 days of COVID-19 exposure.

**Design:** This randomized, double-blind, placebo-controlled trial across the United States (PROTECT-EHC) enrolled in 2020-2021. Participants were followed for 60 days.

**Setting:** Describe the study setting to assist readers to determine the applicability of the report to other circumstances, for example, multicenter, population-based, primary care or referral center(s), etc.

**Participants:** Participants included individuals ≥ 1 year of age with a household contact with a recent (≤ 7 days) diagnosis of COVID-19. 182 participants were enrolled and randomized during the study period.

**Intervention:** Participants were randomized to receive daily oral LGG or microcrystalline cellulose placebo for 28 days.

**Main Outcomes and Measures:** The primary outcome was development of symptoms within 28 days of exposure to a COVID-19-infected household contact. Stool was collected to evaluate for changes in microbiome structure.

**Results:** 182 participants were enrolled and randomized during the study period. Intention-to-treat analysis showed that participants randomized to LGG were less likely to develop symptoms versus those randomized to placebo (26.4% vs. 42.9%, p=0.02). Further, LGG was associated with a statistically significant reduction in COVID-19 diagnosis (log rank p=0.049) via time-to-event analysis. Overall incidence of COVID-19 diagnosis did not significantly differ between LGG and placebo groups (8.8% vs. 15.4%, p=0.17). LGG was well-tolerated with no increased side effects versus placebo. Placebo recipients were more likely to stop the study product, temporarily or permanently, due to symptoms attributed to the study product (5.5% vs. 0%, p = 0.02).

**Conclusions and Relevance:** Our study suggests that LGG is well-tolerated and is associated with prolonged time to development of COVID-19 infection, reduced incidence of symptoms, and changes to gut microbiome structure when used as post-exposure prophylaxis within 7 days after exposure. This preliminary work may inform the approach to prevention of COVID-19, particularly in underdeveloped nations where *Lactobacillus* probiotics have already been utilized to reduce non-COVID sepsis and infectious-morbidity. This study was limited by its remote format, which necessitated a primary endpoint of self-reported symptoms rather than laboratory-confirmed infection; further laboratory-based studies are needed to further define the efficacy of LGG in preventing COVID-19 infection, especially in larger populations and including comparison of pre-exposure vs. post-exposure prophylaxis.

**Trial registration:** ClinicalTrials.gov, NCT04399252, https://clinicaltrials.gov/ct2/show/NCT04399252

**KEY POINTS:** *Question:* Is daily probiotic *Lactobacillus rhamnosus* GG (LGG) effective in preventing development of symptoms of illness compatible with COVID-19 within 28 days of COVID-19 exposure compared to placebo?

*Findings:* In this randomized clinical trial that included 182 participants, the proportion who developed symptoms was 26.4% with LGG versus 42.9% with placebo, a significant difference.

*Meaning:* LGG probiotic may protect against the development of symptoms when used as post-exposure prophylaxis within 7 days after COVID-19 exposure.

## INTRODUCTION

The Coronavirus Disease 2019 (COVID-19) pandemic, caused by severe acute respiratory syndrome coronavirus (SARS-CoV-2) infection, has significantly altered global public health, with over 259 million cases and 5.1 million deaths worldwide as of 29-November-2021.^1^ Despite the advent of highly effective vaccines against SARS-CoV-2, widespread implementation has been slow in areas with limited vaccine availability, with reports estimating that only 3.1% of people in low-income countries have received at least one dose.^2,3^ Vaccine uptake remains limited even in developed nations: only 59% of the U.S. population is fully vaccinated against COVID-19.^1^ Finally, immunity and protection provided by vaccines appears to wane over time.^4^ Thus, additional safe, low-cost, rapidly implementable strategies to address the ongoing COVID-19 pandemic continue to be necessary.

One potential target for intervention is via manipulation of the gut microbiota using probiotics (ingested live bacteria), a well-described strategy to modulate the human immune system and inflammatory responses.^5^ Probiotics have been shown to improve outcomes in a wide variety of infectious presentations including sepsis, ventilator-associated pneumonia, and respiratory tract infections (RTIs).^5-7^ Recent studies suggest that prophylaxis with *Lactobacillus* species specifically can prevent the development of upper and lower RTIs;^8-11^ one large randomized controlled trial of full-term healthy infants randomized to *Lactobacillus* synbiotic vs. placebo showed a 40% reduction in sepsis or death (9.0% vs. 5.4%, p<0.001), including a 34% reduction in lower RTIs (6.1% vs. 4.0%, p=0.002)^8^. These outcomes may be mediated by the effects of probiotics on the immune system and intestinal/lung barrier function via improved intestinal homeostasis, increased regulatory T-cells, normalization of protective mucin production, decreased pro-inflammatory cytokines, modulation of antiviral gene expression, and increased expression of TLRs.^12-16^

These clinical and laboratory reports suggest a potent immunomodulatory role for probiotic therapies in preventing or attenuating respiratory infections, and increasing evidence suggests that gut microbiota affect COVID-19 transmission risk and symptom severity.^17^ Thus, modulation of the gut microbiome via probiotics is a promising strategy for prophylaxis and mitigation of COVID-19. Since March 2020, several trials have launched investigating the benefits of probiotics in both treatment and prevention of COVID-19.^17^ Among commercially-available probiotics, *Lactobacillus rhamnosus* GG (LGG) is particularly encouraging given the success of *Lactobacillus* strains in numerous *in vivo* studies and clinical trials, as discussed above.^8-11^ We therefore conducted a randomized, double-blind, placebo-controlled trial of LGG as post-exposure prophylaxis in exposed household contacts (individuals living with someone recently diagnosed with COVID-19). We hypothesized that LGG prophylaxis would decrease the incidence of symptoms (primary endpoint) and incidence and time to confirmed diagnosis of COVID-19 infection.

## METHODS

### Trial design

Participants were randomized using a permuted block randomization technique to receive LGG or placebo in a 1:1 ratio. Both subjects and study coordinators are blinded to the intervention; the randomization key was generated by the study statistician and only the pharmacist dispensing the study product had access to the key. Due to the ongoing COVID-19 pandemic, the study was designed so that all procedures could be conducted remotely. Study product was delivered by mail, and follow-up was obtained through web-based surveys and telephone calls, with stool samples shipped back to the study center. This research was conducted under Food and Drug Administration Investigational New Drug Application 24777; the research protocol was approved by the Duke University Institutional Review Board, registered on ClinicalTrials.gov (NCT04399252), and was previously published.^18^ All participants provided documented informed consent.

### Subject population and recruitment

Eligibility criteria included: age ≥ one year; exposed household contact (EHC) of someone diagnosed with COVID-19 within the past seven days; willingness to not take any other probiotic while on LGG/placebo; and access to e-mail/internet to complete electronic consent and surveys. Exclusion criteria included: symptoms of COVID-19 at enrollment, including fever, respiratory symptoms (e.g. cough, dyspnea), GI symptoms, anosmia, ageusia; >seven days since index case of household contact had first positive COVID-19 test; taking hydroxychloroquine or remdesivir for any reason; enrolled in a COVID-19 prophylaxis study or receiving COVID-19 prophylaxis as standard of care, including vaccination; any medical condition that would prevent taking oral probiotics or increase risks associated with probiotics; unable to read and follow directions in English or Spanish; living outside of the United States of America; and prisoners and institutionalized individuals. Participants were recruited locally via telephone outreach from study coordinators who identified index cases via the Duke University Hospital Epic dashboard or nationally via flyers, advertisements, social media platforms (https://www.facebook.com/protectehc/), or our study website (https://www.protect-ehc.org/). After electronic consent and randomization, product was dispensed to participants via Federal Express overnight delivery.

### Interventions

Participants took LGG or placebo once daily for 28 days starting from receipt of the blinded shipped study package (age <five, one capsule daily, age ≥five, two capsules daily). LGG capsules, made by Culturelle (DSM), contained ten billion colony forming units of *Lactobacillus rhamnosus* GG (ATCC 53103). The placebo capsules (DSM) contained 325 mg of microcrystalline cellulose, a food additive commonly used as a bulking agent in food preparation and vitamin supplements, and as a placebo in probiotic studies.^19-21^ Both products and their foil packaging were visually indistinguishable.

### Data Collection

Data on demographics, medical history, household risks, and infection details of index patient were collected remotely upon enrollment via REDCap, an electronic platform that supports secure data capture for research.^22^ Data on medications, adherence, COVID-19 exposures, symptoms, adverse events, and COVID-19-related events were collected throughout the study up to day 60. Participants who reported symptoms were queried for laboratory-confirmed COVID-19 infection via electronic health record review, surveys, and phone calls. Subjects self-collected stool using OMNIgene-gut collection kits, which were returned via mail for sequencing analysis.

### Outcomes

The primary endpoint was the development of symptoms, including fever/chills, headache, muscle aches, runny nose, sore throat, cough, shortness of breath, nausea or vomiting, diarrhea, stomach upset or pain, excessive bloating or gas, constipation, loss of sense of smell, loss of sense of taste, rash, painful toes, or other symptoms as reported by participants. Secondary endpoints included: time to COVID-19 diagnosis; incidence of COVID-19 diagnosis, severity of symptoms; and duration of symptoms. In participants who reported diagnosis of COVID-19, we reviewed medical records for laboratory confirmation of the diagnosis as well as complications (e.g., need for hospitalization, intubation, mortality), when available. We investigated the incidence of these events through day 28 and through day 60.

### Sequencing Analysis

DNA from stool samples was extracted using the Qiagen PowerSoil DNA kit, and the V4 region of the 16S rRNA gene was PCR amplified and sequenced on the Illumina MiSeq platform as previously described.^23^ After demultiplexing, DADA2 was used for quality control and to generate a count table,^24^ and taxonomy was assigned using the Silva v138.1 database. Data analysis was performed using the R programming suite packages phyloseq and ggplot2. PERMANOVA testing was performed to assess for statistical significance in principal coordinates analyses. The sequencing data for this project is available at SRAxxxxxxx (to be made available once accepted).

### Statistical Analysis

Analyses were conducted via intention-to-treat (ITT) methodology, including all participants who were enrolled and randomized. Additionally, we performed pre-specified analysis with modified ITT methodology including all enrolled and randomized participants who confirmed physical receipt of the study product (mITTrt) as well as a pre-specified analysis that included enrolled and randomized participants who confirmed physical receipt of the study product and remained symptom free at the time of study product receipt (mITTasymptomatic). All analyses were performed in SAS 9.4 (SAS Institute, Cary, NC). Chi-squared tests were employed to test the differences in COVID-19 symptoms, laboratory-confirmed infections, and other categorical variables between the LGG and placebo arms. Student’s t-tests were used to compare continuous variables such as symptom duration and adherence. Kaplan-Meier curves were constructed, and log-rank tests used to test the univariable differences in time-to-infection/symptom outcomes. Logistic regression modeling of day 28 symptoms was performed in mITTrt cohort to adjust for the potential confounding caused by age and smoking status. Pre-study sample size was calculated assuming an attack rate of 10.5% in household contacts based on CDC reports.^25^ With 1076 participants (538 per arm), the chi-squared test with 1-sided alpha=5% would have 80% power to detect a 40% reduction (estimated from data showing 30-50% reduction in respiratory infections with LGG.^8,26,27^) in the attack rate of COVID-19, from 10.5 to 6.3%.

## RESULTS

### Participant characteristics

Enrollment was stopped early on June 2, 2021, after the study team noted changes in recruitment patterns such that most individuals approached for the study had already been vaccinated and were therefore ineligible. During this period, 182 participants were enrolled and randomized (ITT). Of these, 135 confirmed that they physically received and started the study product and were considered to have received therapy (mITTrt); the other 47 participants did not respond to repeated queries. Of those 135, 31 participants reported development of symptoms prior to receiving study product; 104 remained asymptomatic at initiation of therapy (mITTasymptomatic) (Figure 1). The demographic characteristics of the ITT participants are displayed in Table 1; demographics of mITTrt and mITTasymptomatic analyses are available in Supplementary Tables 1a and 1b. Groups were evenly balanced other than increased prevalence of smoking (14.3% vs. 4.4%) and hypertension in the placebo group (18.7% vs. 5.5%). There were no differences in employment in healthcare, recent visits to healthcare facilities, use of’ probiotics or antibiotics prior to the start of the study, frequency of mask wearing, social distancing, and handwashing between groups (Supplementary Table 2a,b,c, all p > 0.05).

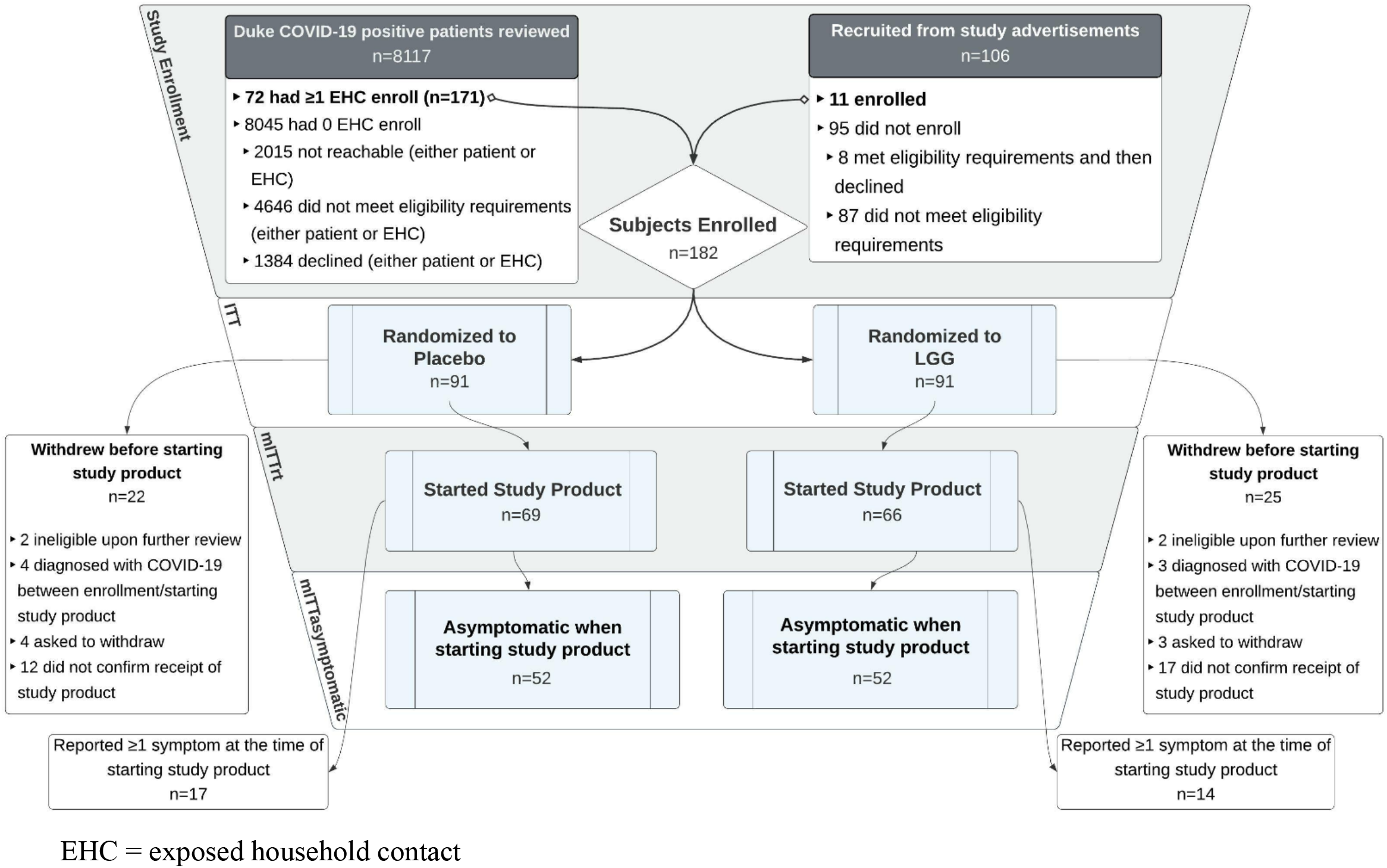

**Table 1.** Demographic and Clinical Characteristics of Participants at Baseline, ITT analysis.

|  | **LGG** | **Placebo** | **All**<br>**Participants** |
| --- | --- | --- | --- |
|  | **N=91**<br>**(50%)** | **N=91**<br>**(50%)** | **N=182**<br>**(100%)** |
| **Age group – no. (%)** |  |  |  |
| - <18 | 25 (27.5%) | 16 (17.6%) | 41 (22.5%) |
| - 18-64 | 64 (70.3%) | 67 (73.6%) | 131 (72%) |
| - ≥65 | 2 (2.2%) | 8 (8.8%) | 10 (5.5%) |
| **Female sex – no. (%)** | 60 (65.9%) | 55 (60.4%) | 115 (63.2%) |
| **Race – no. (%)** |  |  |  |
| - White | 55 (60.4%) | 66 (72.5%) | 121 (66.5%) |
| - Black | 18 (19.8%) | 17 (18.7%) | 35 (19.2%) |
| - Asian | 2 (2.2%) | 2 (2.2%) | 4 (2.2%) |
| - Other | 11 (12.1%) | 4 (4.4%) | 15 (8.2%) |
| - More Than One | 4 (4.4%) | 1 (1.1%) | 5 (2.7%) |
|  | **N=91**<br>**(50%)** | **N=91**<br>**(50%)** | **N=182**<br>**(100%)** |
| - Not Reported | 1 (1.1%) | 1 (1.1%) | 2 (1.1%) |
| **Hispanic Ethnicity – no. (%)** | 14 (15.4%) | 12 (13.2%) | 26 (14.3%) |
| **Comorbid Conditions – no. (%)** |  |  |  |
| - Current smoker | 4 (4.4%) | 13 (14.3%) | 17 (9.3%) |
| - Lung disease | 0 (0%) | 2 (2.2%) | 2 (1.1%) |
| - Allergies | 13 (14.3%) | 25 (27.5%) | 38 (20.9%) |
| - Cancer | 1 (1.1%) | 4 (4.4%) | 5 (2.7%) |
| - Hypertension | 5 (5.5%) | 17 (18.7%) | 22 (12.1%) |
| - Diabetes | 2 (2.2%) | 5 (5.5%) | 7 (3.8%) |
| - Heart disease/stroke | 2 (2.2%) | 5 (5.5%) | 7 (3.8%) |
| - Liver disease | 0 (0%) | 1 (1.1%) | 1 (0.5%) |
| - Currently pregnant | 2 (2.2%) | 1 (1.1%) | 3 (1.6%) |
| **Antibiotic Use within past 30 days – no. (%)** | 4 (4.4%) | 6 (6.6%) | 10 (5.5%) |
| **Probiotic Use within past 30 days – no. (%)** | 5 (5.5%) | 6 (6.6%) | 11 (6%) |
| **Days from exposure to enrollment – median (IQR)** | 2 (1 - 3) | 2 (1 - 2) | 2 (1 - 2) |
| **Days from exposure to study product start – median (IQR)** | 3.5 (2 - 5) | 3 (2 - 4) | 3 (2 - 4) |
| - 0 | 30 (33%) | 35 (38.5%) | 65 (35.7%) |
| - 1 | 11 (12.1%) | 10 (11%) | 21 (11.5%) |
|  | **N=91**<br>**(50%)** | **N=91**<br>**(50%)** | **N=182**<br>**(100%)** |
| - 2 | 18 (19.8%) | 22 (24.2%) | 40 (22%) |
| - 3 | 3 (3.3%) | 5 (5.5%) | 8 (4.4%) |
| - 4 | 12 (13.2%) | 11 (12.1%) | 23 (12.6%) |
| - Unknown | 17 (18.7%) | 8 (8.8%) | 25 (13.7%) |

### COVID-19 symptoms and infection

Participants randomized to LGG were significantly less likely to report any symptoms by day 28 (26.4% vs. 42.9%, p = 0.02, Table 2). No participants reported new symptoms after day 28. Participants receiving LGG had significantly prolonged time to onset of symptoms (log rank p = 0.006, Figure 2a). There was no difference in the proportion of participants who reported specific symptoms in any of the analysis subgroups, though placebo recipients were more likely experience moderate to severe changes in taste perception (5.5% vs. 0%, p = 0.02, Supplementary Table 3).

**Table 2.** Outcomes of LGG Therapy for Postexposure Prophylaxis against COVID-19 at D28.

|  | **LGG** | **Placebo** | **All**<br>**Participants** |  |
| --- | --- | --- | --- | --- |
| **ITT** | **N=91**<br>**(50%)** | **N=91**<br>**(50%)** | **N=182**<br>**(100%)** | **P-Value** |
| **Any symptoms** – no. (%) | 24 (26.4%) | 39 (42.9%) | 63 (34.6%) | 0.02 |
| **Any moderate/severe symptoms** – no. (%) | 16 (17.6%) | 24 (26.4%) | 40 (22%) | 0.15 |
| **Symptom duration** – median (IQR) | 8 (4 - 17) | 11 (4 - 22) | 10 (4 - 21) | 0.37 |
| **Reported COVID-19 Diagnosis** – no. (%) | 8 (8.8%) | 14 (15.4%) | 22 (12.1%) | 0.17 |
| **mITT<sub>TrT</sub>** | **N=66**<br>**(48.9%)** | **N=69**<br>**(51.1%)** | **N=135**<br>**(100%)** |  |
| **Any symptoms** – no. (%) | 24 (36.4%) | 39 (56.5%) | 63 (46.7%) | 0.02 |
| **Any moderate/severe symptoms** – no. (%) | 16 (24.2%) | 24 (34.8%) | 40 (29.6%) | 0.18 |
| **Symptom duration** – median (IQR) | 9.5 (6 - 18) | 12 (4 - 27) | 11 (5.5 - 22) | 0.38 |
| **Reported COVID-19 Diagnosis** – no. (%) | 6 (9.1%) | 13 (18.8%) | 19 (14.1%) | 0.10 |
| **mITT<sub>Asymptomatic</sub>** | **N=52**<br>**(50%)** | **N=52**<br>**(50%)** | **N=104**<br>**(100%)** |  |
| **Any symptoms** – no. (%) | 14 (26.9%) | 25 (48.1%) | 39 (37.5%) | 0.03 |
| **Any moderate/severe symptoms** – no. (%) | 10 (19.2%) | 15 (28.8%) | 25 (24%) | 0.25 |
| **Symptom duration** – median (IQR) | 6.5 (5 - 9.5) | 10 (4 - 18) | 9 (4 - 12) | 0.11 |
| **Reported COVID-19 Diagnosis** – no. (%) | 2 (3.8%) | 7 (13.5%) | 9 (8.7%) | 0.08 |

EHC = exposed household contact

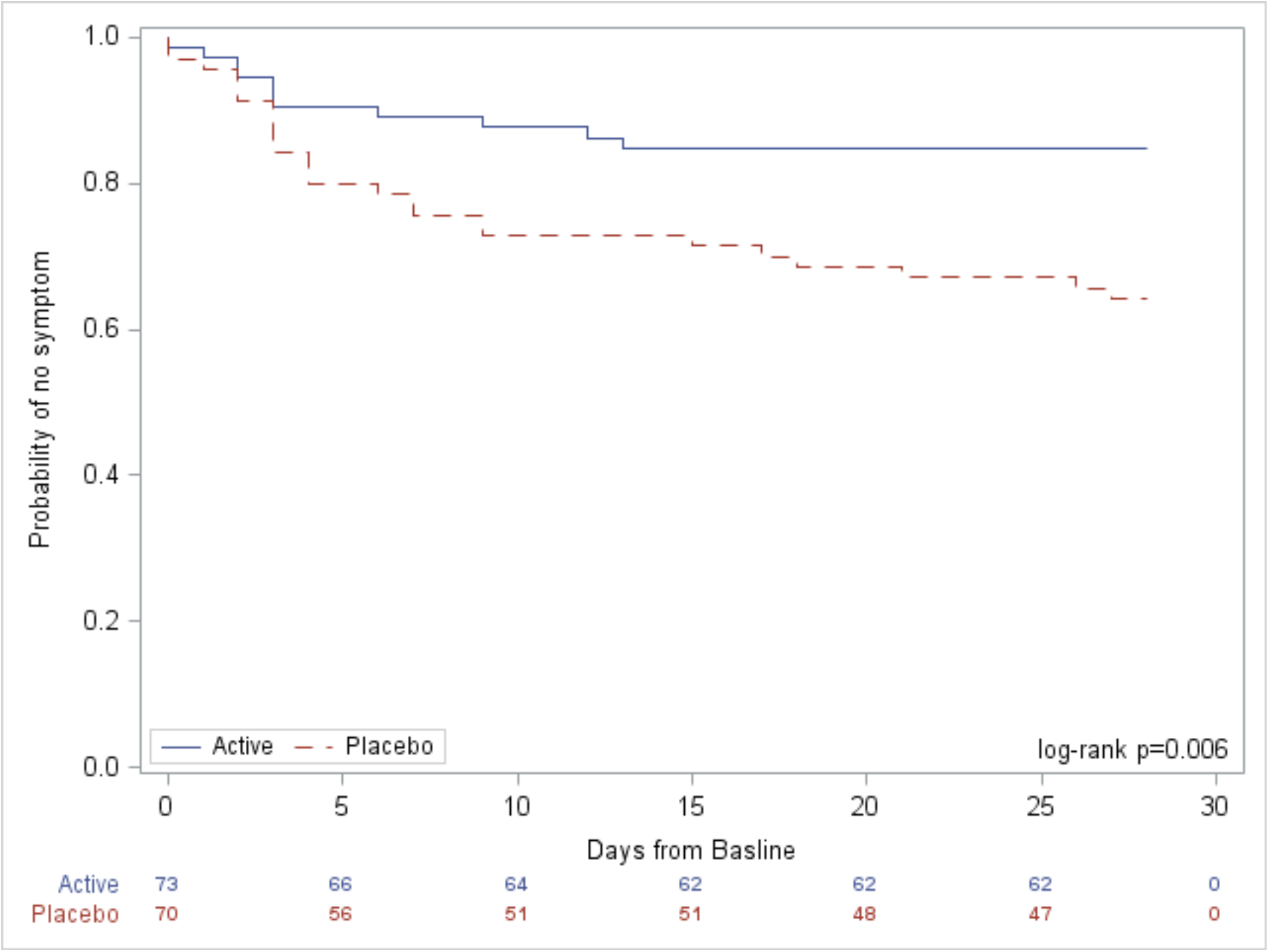
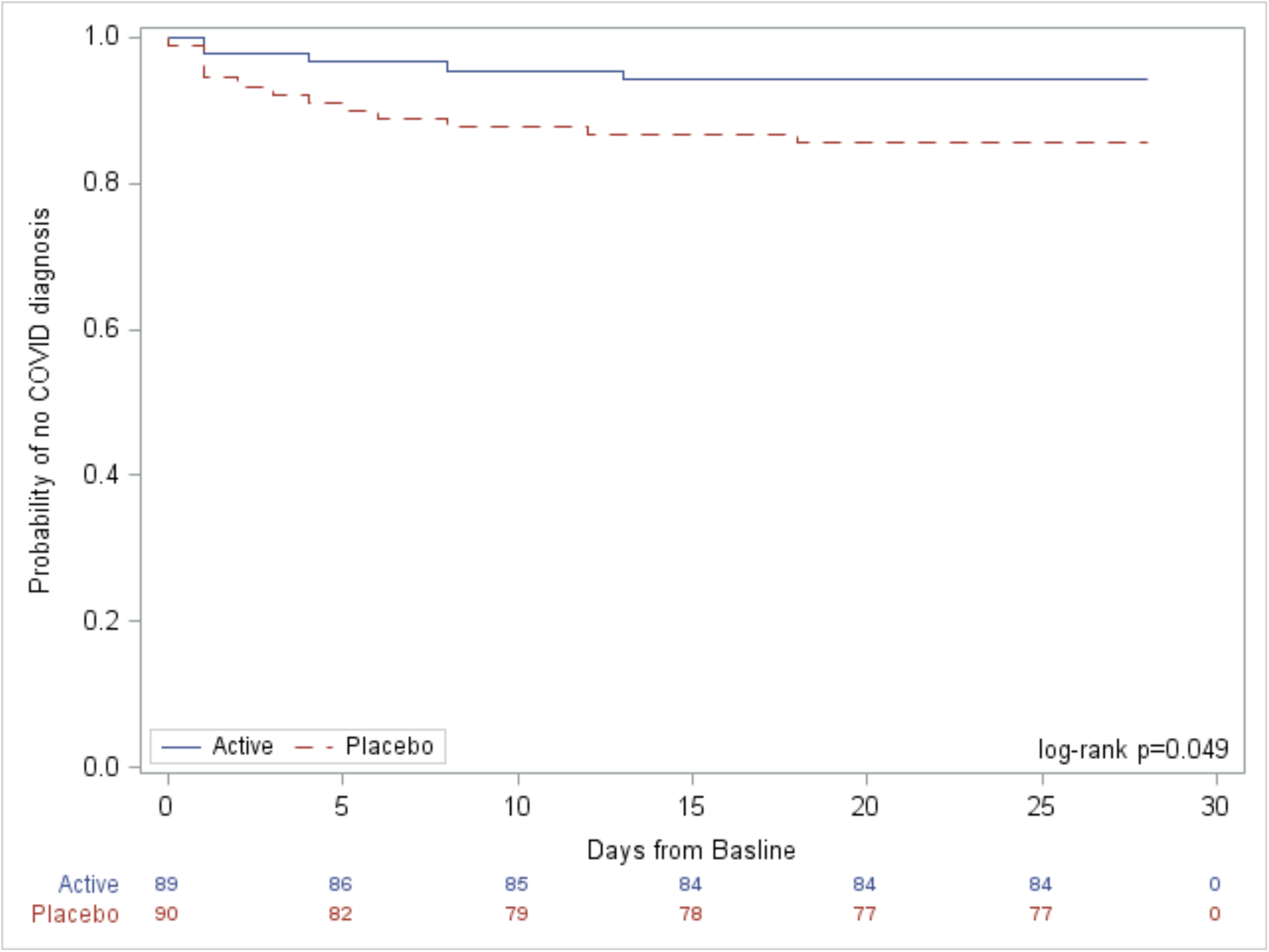

Of 77 symptomatic participants during the study period, 47 underwent testing under the care of their medical provider and 22 had laboratory confirmed COVID-19. Of these, 16 diagnoses were made by PCR testing and confirmed by electronic medical record review, and six were self-reported by participants after laboratory testing. While there was a trend to decreased COVID-19 incidence in participants randomized to LGG, this difference was not statistically significant (8.8% vs. 15.4%, p = 0.17, Table 2); however, time to COVID-19 diagnosis was significantly prolonged for LGG recipients (log rank p = 0.049, Figure 2b). There were no hospitalizations or deaths among any participants. Similar findings were observed in the modified ITT analyses (Table 2, Supplementary Figures 1 and 2), including a trend to a decreased incidence of COVID-19 in subjects not reporting symptoms at initiation of treatment (mITTasymptomatic, 3.8% vs. 13.5%, p=0.08).

### Sensitivity Analyses

Univariate sensitivity analysis by sex revealed no differences in development of COVID-19 symptoms or laboratory-confirmed infection (Supplementary Table 4). Univariate sensitivity analysis by age showed older participants were significantly more likely to report symptoms (age <18, 14.6% vs. age 18-64, 38.9%, vs. age ≥65, 60.0%, p = 0.004) and have laboratory-confirmed infection (age <18, 12.2% vs. age 18-64, 9.9%, vs. age ≥65, 40.0%, p = 0.02, Supplementary Table 5) at day 28. Multivariate logistic regression modeling revealed that age <18 was associated with significantly lower odds of developing symptoms by day 28 compared to the 18-64 age group (OR 0.29, 95%CI 0.1-0.82, p = 0.02); current smoking status was not associated with development of symptoms (OR 0.99, 95%CI 0.33 - 2.98, p = 0.98, Supplementary Table 6).

### Microbiome Analyses

A total of 261 stool samples were received from 106 participants (all in the mITTrt group), with 85 day 7 samples and 69 day 28 samples. participants who received LGG had a significantly greater abundance of *L. rhamnosus* compared to participants who received placebo (Figure 3a). Although there was no difference in the α-diversity between participants who received placebo or probiotic (data not shown), there was a significant difference in the overall structure of the stool microbiota (i.e., β-diversity) (Figure 3b; p = 0.0005). Additionally, the presence of symptoms and a COVID-19 diagnosis significantly affected β-diversity, as did interactions between the treatment group, symptoms, and COVID-19 diagnosis.

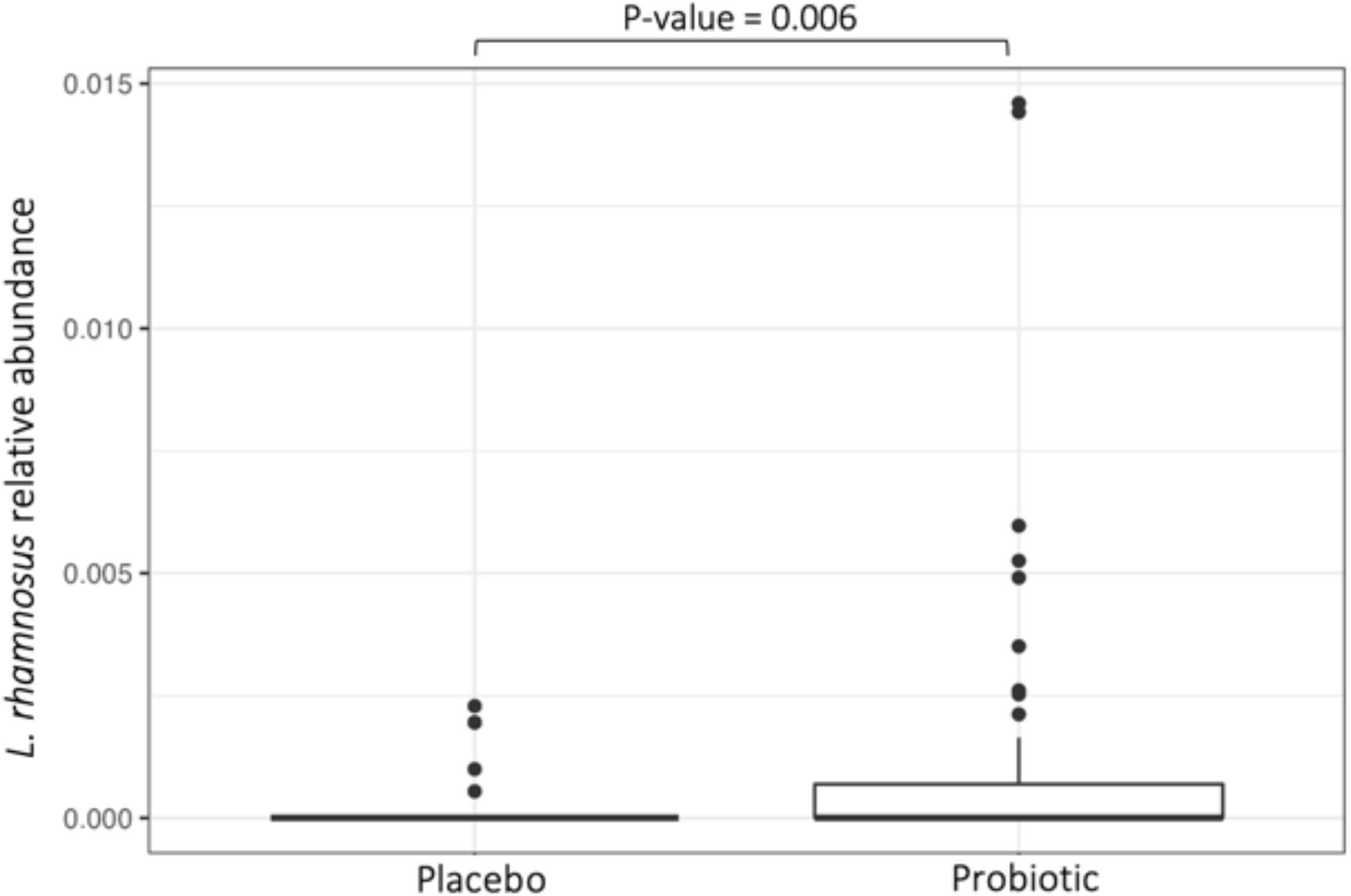
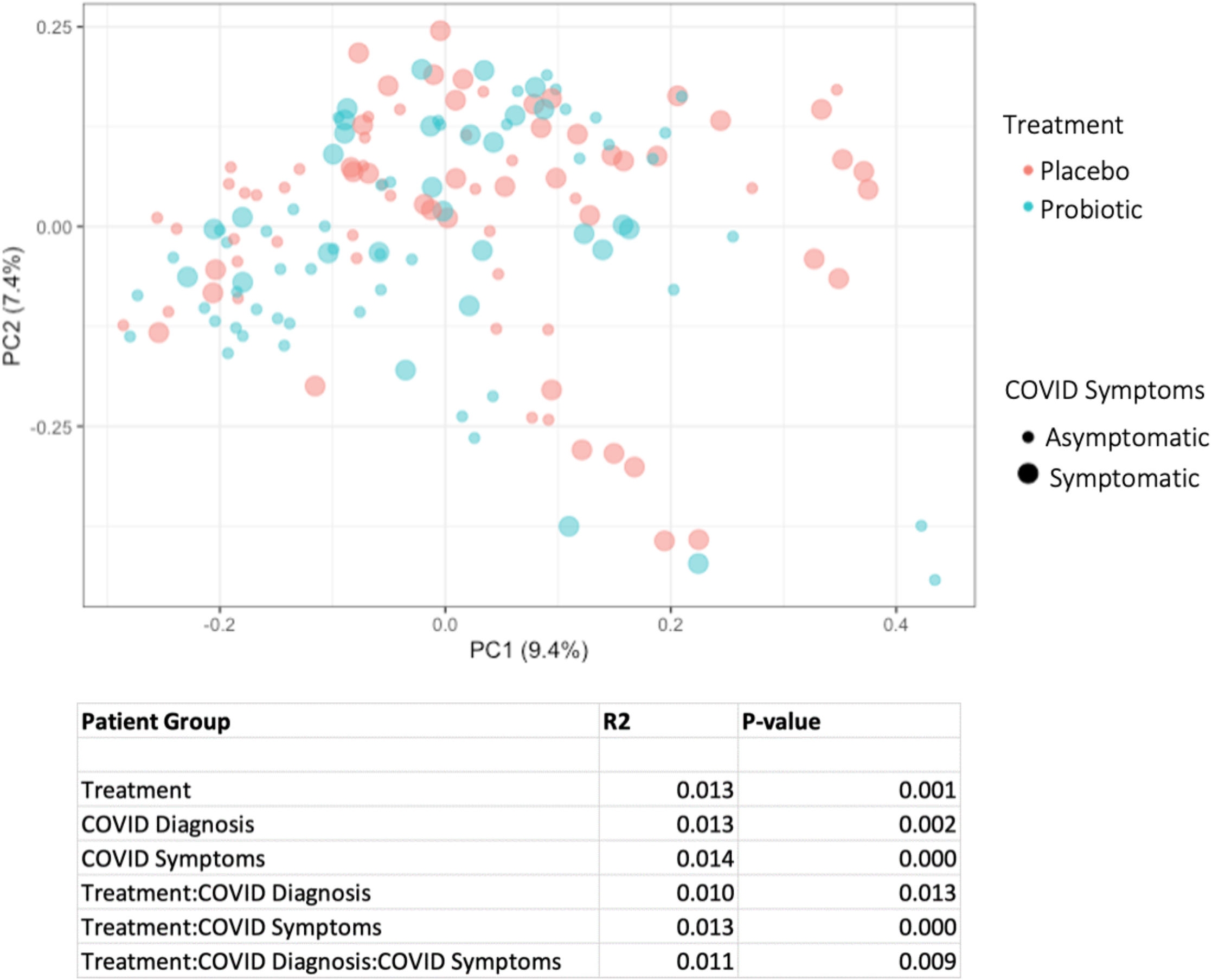

### Adherence and Safety

Of 110 participants who reported at least one adherence time point, median adherence did not differ between LGG and placebo groups (median 100%, IQR 93-100% vs. 100%, IQR 93-100%, p = 0.82, Supplementary Table 7). Participants were unable to guess their randomization arm, suggesting that blinding was maintained. There was no significant difference in proportion of participants who attributed symptoms they experienced to LGG vs. placebo (8.8% vs. 23.1%, p = 0.32), though placebo recipients were more likely to stop the study product, temporarily or permanently, due to symptoms attributed to the study product (5.5% vs. 0%, p = 0.02, Supplementary Table 7). These findings held true in mITTrt and mITTasymptomatic analyses (Supplementary Table 8a, 8b).

## DISCUSSION

In this randomized, double-blind, placebo-controlled trial, we investigated the efficacy of the probiotic LGG as post-exposure prophylaxis against COVID-19. Participants randomized to LGG had fewer symptoms and prolonged time to development of COVID-19 compared to those receiving placebo; this finding held true in all three of our analyses (ITT, mITTrt, and mITTasymptomatic). Interestingly, the data suggest that placebo recipients were more likely to experience moderate to severe changes in taste perception– a relatively specific symptom for COVID-19. A similar trend was observed for changes in smell perception but did not reach statistical significance. While placebo recipients were more likely to stop using study product, the observed rate of gastrointestinal side effects in this study is comparable to other probiotic intervention studies using microcrystalline cellulose as a placebo, which note a 15-20% incidence of gastrointestinal side effects.^19,28,29^ Microbiome analyses confirmed that *L. rhamnosus* abundance was significantly increased in participants who received LGG compared to placebo, suggesting that participants were adherent with study therapy and that microbial community structure differentiated in response to probiotic treatment.

Our study has several limitations. First, we were limited by a smaller-than-expected sample size due to difficulty with recruitment during concurrent vaccine rollout, which increasingly limited the eligible population and our statistical power. Given the high transmissibility of newer viral strains and potential for waning vaccine efficacy, future studies may consider including vaccinated individuals, especially as data suggest that probiotic administration improves vaccination efficacy against other viral pathogens, such as influenza.^30^ Further, while allocation was blinded and randomized in a 1:1 fashion, participants in the placebo group had a small increased incidence of current smoking and hypertension at baseline, which are potential risk factors for development of COVID-19 disease; however, smoking was not associated with development of symptoms in our study. Additionally, LGG and other probiotics may be associated with gastrointestinal side effects, potentially confounding our measurement of symptoms, although fewer GI side effects were noted in the probiotic group. Another limitation was the remote format, wherein the primary endpoint was self-reported symptoms rather than laboratory-confirmed infection; participants had inconsistent access to laboratory testing, with only 61% of symptomatic participants ultimately undergoing testing.

In conclusion, COVID-19 continues to pose a unique and novel challenge to global health.^5^ We conducted the first double-blinded, randomized, placebo-controlled trial to evaluate the effect of prophylaxis with probiotic LGG on development of COVID-19 symptoms in exposed household contacts. While limited in sample size, our study suggests that LGG is well-tolerated and is associated with prolonged time to development of COVID-19 infection, reduced symptomatic disease, and changes to gut microbiome structure. Further investigation of LGG probiotic intervention in larger randomized controlled trials is warranted, including comparison of pre-exposure vs. post-exposure prophylaxis with LGG probiotic in high-risk populations. Our results lend credence to the notion that our symbiotic microbes can be valuable partners in the fight against COVID-19 and potentially other future pandemic diseases.

## Data Availability

Microbiome sequencing data will be made available in the Sequence Read Archive (upon manuscript acceptance). We do not plan to share individual participant data.

