## Supplemental files for "Daily Lactobacillus Probiotic versus Placebo in COVID-19-Exposed Household Contacts (PROTECT-EHC): A Randomized Clinical Trial"

**Supplementary Appendix**

**Figure S1a/b.** Kaplan-Meier curve of time to event mITTrt analysis.

a. Time to any symptoms (N=102, event=35, log rank p=0.01). Participants receiving LGG had prolonged time to onset of any symptoms. Participants who had any symptoms at study start were excluded. Curves were right censored at D28.

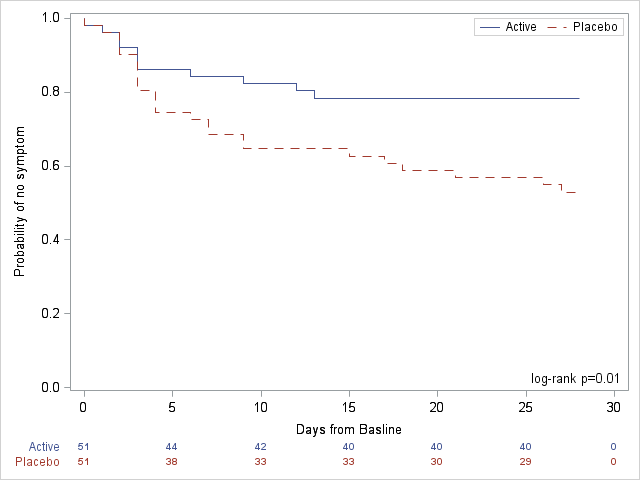

b. Time to reported laboratory-confirmed infection (N=133, event=16, log rank p=0.04). Participants who reported laboratory-confirmed infection at study start were excluded. Curves were right censored at D28.

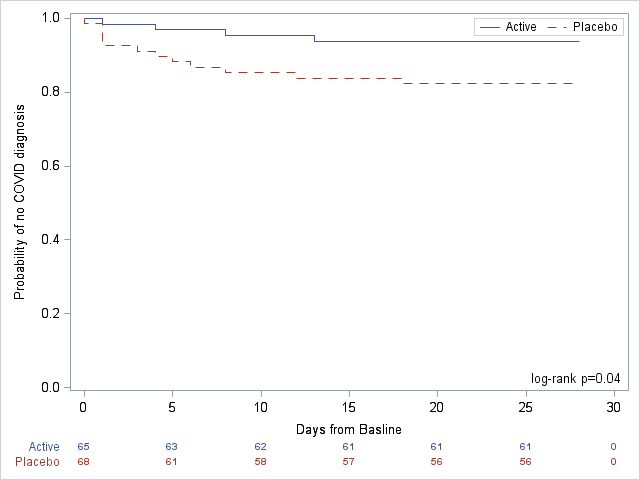

**Figure S2a/b.** Kaplan-Meier curve of time to event (a. symptoms, b. laboratory-confirmed infection), mITTasymptomatic analysis.

a. Time to any symptoms (N=101, event=34, log rank p=0.02). Participants receiving LGG had prolonged time to onset of any symptoms. Participants who had any symptoms at study start were excluded. Curves were right censored at D28.

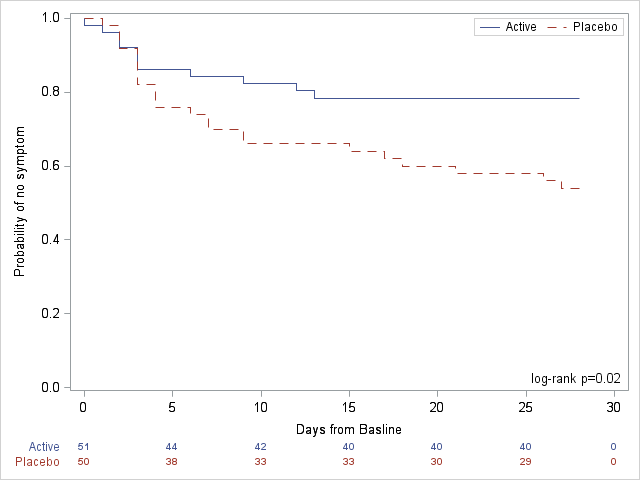

b. Time to reported laboratory-confirmed infection (N=103, event=1-, log rank p=0.05). Participants who reported laboratory-confirmed infection at study start were excluded. Curves were right censored at D28.

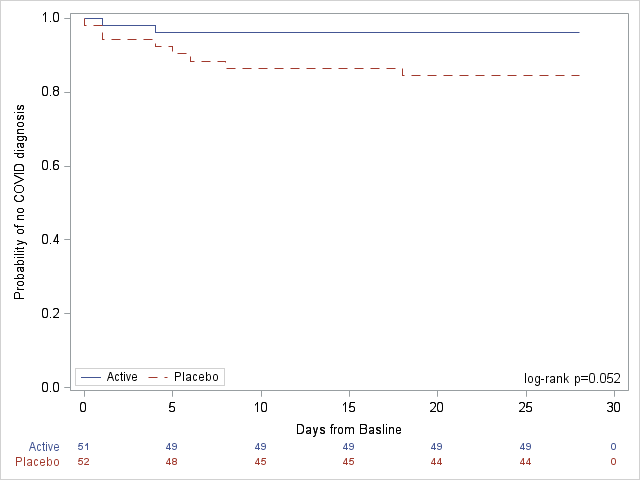

**Table S1a.** Demographic and Clinical Characteristics of Participants at Baseline, mITTrt analysis.

|  | **LGG** | **Placebo** | **All Participants** |
| --- | --- | --- | --- |
|  | **N=66**  **(48.9%)** | **N=69**  **(51.1%)** | **N=135**  **(100%)** |
| **CTGov Age group – no. (%)** |  |  |  |
| - 18-64 | 47 (71.2%) | 52 (75.4%) | 99 (73.3%) |
| - <18 | 17 (25.8%) | 10 (14.5%) | 27 (20%) |
| - >=65 | 2 (3%) | 7 (10.1%) | 9 (6.7%) |
| **Female sex** – no. (%) | 43 (65.2%) | 44 (63.8%) | 87 (64.4%) |
| **Race** – no. (%) | | | |
| - White | 46 (69.7%) | 53 (76.8%) | 99 (73.3%) |
| - Black | 10 (15.2%) | 11 (15.9%) | 21 (15.6%) |
| - Other | 6 (9.1%) | 2 (2.9%) | 8 (5.9%) |
| - More Than One | 3 (4.5%) | 1 (1.4%) | 4 (3%) |
| - Asian | 1 (1.5%) | 2 (2.9%) | 3 (2.2%) |
| **Hispanic Ethnicity** – no. (%) | 8 (12.1%) | 7 (10.1%) | 15 (11.1%) |
| **Comorbid Conditions** – no. (%) | | | |
| - Current smoker | 4 (6.1%) | 12 (17.4%) | 16 (11.9%) |
| - Cancer | 0 (0%) | 4 (5.8%) | 4 (3%) |
| - Lung disease | 0 (0%) | 2 (2.9%) | 2 (1.5%) |
| - Allergies | 13 (19.7%) | 22 (31.9%) | 35 (25.9%) |
| - Diabetes | 2 (3%) | 2 (2.9%) | 4 (3%) |
| - Hypertension | 5 (7.6%) | 14 (20.3%) | 19 (14.1%) |
| - Heart disease/stroke | 2 (3%) | 2 (2.9%) | 4 (3%) |
| - Liver disease | 0 (0%) | 1 (1.4%) | 1 (0.7%) |
| - Currently pregnant | 2 (3%) | 1 (1.4%) | 3 (2.2%) |
| **Antibiotic Use within past 30 days** – no. (%) | 2 (3%) | 5 (7.2%) | 7 (5.2%) |
| **Probiotic Use within past 30 days** – no. (%) | 5 (7.6%) | 6 (8.7%) | 11 (8.1%) |
| **Days from exposure to enrollment** – median (IQR) | 2 (1 - 2) | 2 (1 - 3) | 2 (1 - 2) |
| **Days from exposure to study product start** – median (IQR) | 3.5 (2 - 5) | 3 (2 - 4) | 3 (2 - 4) |
| **Household Risk Score** | | | |
| - 0 | 25 (37.9%) | 30 (43.5%) | 55 (40.7%) |
| - 1 | 8 (12.1%) | 10 (14.5%) | 18 (13.3%) |
| - 2 | 15 (22.7%) | 16 (23.2%) | 31 (23%) |
| - 3 | 3 (4.5%) | 5 (7.2%) | 8 (5.9%) |
| - 4 | 10 (15.2%) | 8 (11.6%) | 18 (13.3%) |
| - Unknown | 5 (7.6%) | 0 (0%) | 5 (3.7%) |

**Table S1b.** Demographic and Clinical Characteristics of Participants at Baseline, mITTasymptomatic analysis.

|  | **LGG** | **Placebo** | **All Participants** |
| --- | --- | --- | --- |
|  | **N=52 (50%)** | **N=52 (50%)** | **N=104 (100%)** |
| **CTGov Age group – no. (%)** | | | |
| - 18-64 | 35 (67.3%) | 37 (71.2%) | 72 (69.2%) |
| - <18 | 15 (28.8%) | 10 (19.2%) | 25 (24%) |
| - >=65 | 2 (3.8%) | 5 (9.6%) | 7 (6.7%) |
| **Female sex** – no. (%) | 33 (63.5%) | 30 (57.7%) | 63 (60.6%) |
| **Race** – no. (%) | | | |
| - White | 35 (67.3%) | 40 (76.9%) | 75 (72.1%) |
| - Black | 9 (17.3%) | 8 (15.4%) | 17 (16.3%) |
| - Other | 6 (11.5%) | 2 (3.8%) | 8 (7.7%) |
| - More Than One | 2 (3.8%) | 1 (1.9%) | 3 (2.9%) |
| - Asian | 0 (0%) | 1 (1.9%) | 1 (1%) |
| **Hispanic Ethnicity** – no. (%) | 8 (15.4%) | 6 (11.5%) | 14 (13.5%) |
| **Comorbid Conditions** – no. (%) | | | |
| - Current smoker | 4 (7.7%) | 10 (19.2%) | 14 (13.5%) |
| - Cancer | 0 (0%) | 3 (5.8%) | 3 (2.9%) |
| - Lung disease | 0 (0%) | 1 (1.9%) | 1 (1%) |
| - Allergies | 8 (15.4%) | 14 (26.9%) | 22 (21.2%) |
| - Diabetes | 1 (1.9%) | 2 (3.8%) | 3 (2.9%) |
| - Hypertension | 2 (3.8%) | 10 (19.2%) | 12 (11.5%) |
| - Heart disease/stroke | 2 (3.8%) | 2 (3.8%) | 4 (3.8%) |
| - Liver disease | 0 (0%) | 1 (1.9%) | 1 (1%) |
| - Currently pregnant | 2 (3.8%) | 0 (0%) | 2 (1.9%) |
| **Antibiotic Use within past 30 days** – no. (%) | 1 (1.9%) | 2 (3.8%) | 3 (2.9%) |
| **Probiotic Use within past 30 days** – no. (%) | 4 (7.7%) | 4 (7.7%) | 8 (7.7%) |
| **Days from exposure to enrollment** – median (IQR) | 2 (1 - 2) | 1 (1 - 2) | 2 (1 - 2) |
| **Days from exposure to study product start** – median (IQR) | 3 (2 - 4) | 3 (2 - 4) | 3 (2 - 4) |
| **Household Risk Score** | | | |
| - 0 | 19 (36.5%) | 26 (50%) | 45 (43.3%) |
| - 1 | 7 (13.5%) | 8 (15.4%) | 15 (14.4%) |
| - 2 | 13 (25%) | 9 (17.3%) | 22 (21.2%) |
| - 3 | 2 (3.8%) | 4 (7.7%) | 6 (5.8%) |
| - 4 | 6 (11.5%) | 5 (9.6%) | 11 (10.6%) |
| - Unknown | 5 (9.6%) | 0 (0%) | 5 (4.8%) |

**Table S2a.** COVID-19 exposure risk factors, ITT analysis.

|  | **LGG** | **Placebo** | **All Participants** |  |
| --- | --- | --- | --- | --- |
|  | **N=91**  **(50%)** | **N=91**  **(50%)** | **N=182**  **(100%)** | **P-Value** |
| **Household size** – no. (%) | | | | |
| - 2 | 15 (16.5%) | 27 (29.7%) | 42 (23.1%) | 0.38 |
| - 3 | 15 (16.5%) | 11 (12.1%) | 26 (14.3%) |  |
| - 4 | 15 (16.5%) | 22 (24.2%) | 37 (20.3%) |  |
| - 5 | 14 (15.4%) | 12 (13.2%) | 26 (14.3%) |  |
| - ≥6 | 13 (14.3%) | 9 (9.9%) | 22 (12.1%) |  |
| - Not reported | 19 (20.9%) | 10 (11%) | 29 (15.9%) |  |
| **COVID-positive contacts in household** – no. (%) | | | | |
| - 1 | 37 (40.7%) | 36 (39.6%) | 73 (40.1%) | 0.49 |
| - 2 | 10 (11%) | 8 (8.8%) | 18 (9.9%) |  |
| - 3 | 2 (2.2%) | 3 (3.3%) | 5 (2.7%) |  |
| - 4 | 0 (0%) | 2 (2.2%) | 2 (1.1%) |  |
| - Not reported | 42 (46.2%) | 42 (46.2%) | 84 (46.2%) |  |
| **Close contact with COVID-positive person outside home within past week** – no (%) | 10 (11%) | 14 (15.4%) | 24 (13.2%) | 0.58 |
| **Participation in large social gatherings** **within past week** – no. (%) | 7 (7.7%) | 10 (11%) | 17 (9.3%) | 0.66 |
| **Employment outside of home** – no. (%) | 30 (33%) | 41 (45.1%) | 71 (39%) | 0.30 |
| **Employment at a healthcare facility** – no. (%) | 8 (8.8%) | 17 (18.7%) | 25 (13.7%) | 0.11 |
| **Visit to healthcare facility within past week** – no. (%) | 16 (17.6%) | 24 (26.4%) | 40 (22%) | 0.32 |
| **Adherence to mask wearing** – no. (%) |  |  |  |  |
| - Always | 51 (56%) | 61 (67%) | 112 (61.5%) | 0.71 |
| - Often | 13 (14.3%) | 16 (17.6%) | 29 (15.9%) |  |
| - Sometimes | 3 (3.3%) | 1 (1.1%) | 4 (2.2%) |  |
| - Rarely | 0 (0%) | 0 (0%) | 0 (0%) |  |
| - Never | 1 (1.1%) | 1 (1.1%) | 2 (1.1%) |  |
| - Not reported | 23 (25.3%) | 12 (13.1%) | 35 (19.2%) |  |
| **Adherence to social distancing** – no. (%) |  |  |  |  |
| - Always | 37 (40.7%) | 54 (59.3%) | 91 (50%) | 0.12 |
| - Often | 29 (31.9%) | 21 (23.1%) | 50 (27.5%) |  |
| - Sometimes | 2 (2.2%) | 4 (4.4%) | 6 (3.3%) |  |
| - Rarely | 0 (0%) | 0 (0%) | 0 (0%) |  |
| - Never | 0 (0%) | 0 (0%) | 0 (0%) |  |
| - Not reported | 23 (25.3%) | 12 (13.1%) | 35 (19.2%) |  |
| **Adherence to handwashing** – no. (%) | | | | |
| - Always | 40 (44%) | 58 (63.7%) | 98 (53.8%) | 0.06 |
| - Often | 24 (26.4%) | 14 (15.4%) | 38 (20.9%) |  |
| - Sometimes | 4 (4.4%) | 5 (5.5%) | 9 (4.9%) |  |
| - Rarely | 0 (0%) | 2 (2.2%) | 2 (1.1%) |  |
| - Never | 0 (0%) | 0 (0%) | 0 (0%) |  |
| - Not reported | 23 (25.3%) | 12 (13.1%) | 35 (19.2%) |  |

**Table S2b.** COVID-19 exposure risk factors, mITTrtanalysis.

|  | **LGG** | **Placebo** | **All Participants** |  |
| --- | --- | --- | --- | --- |
|  | **N=66**  **(48.9%)** | **N=69**  **(51.1%)** | **N=135**  **(100%)** | **P-Value** |
| **Household size** – no. (%) | | | | |
| - 2 | 12 (18.2%) | 23 (33.3%) | 35 (25.9%) | 0.47 |
| - 3 | 13 (19.7%) | 11 (15.9%) | 24 (17.8%) |  |
| - 4 | 13 (19.7%) | 17 (24.6%) | 30 (22.2%) |  |
| - 5 | 12 (18.2%) | 11 (15.9%) | 23 (17%) |  |
| - ≥6 | 10 (15.2%) | 5 (7.6%) | 15 (11.1%) |  |
| - Not reported | 6 (9.1%) | 2 (2.9%) | 8 (5.9%) |  |
| **COVID-positive contacts in household** – no. (%) | | | | |
| - 1 | 32 (48.5%) | 29 (42%) | 61 (45.2%) | 0.33 |
| - 2 | 8 (12.1%) | 6 (8.7%) | 14 (10.4%) |  |
| - 3 | 1 (1.5%) | 3 (4.3%) | 4 (3%) |  |
| - 4 | 0 (0%) | 2 (2.9%) | 2 (1.5%) |  |
| - Not reported | 25 (37.9%) | 29 (42%) | 54 (40%) |  |
| **Close contact with COVID-positive person outside home within past week** – no (%) | 10 (15.2%) | 13 (18.8%) | 23 (17%) | 0.72 |
| **Participation in large social gatherings** **within past week** – no. (%) | 6 (9.1%) | 9 (13%) | 15 (11.1%) | 0.62 |
| **Employment outside of home** – no. (%) | 27 (40.9%) | 34 (49.3%) | 61 (45.2%) | 0.57 |
| **Employment at a healthcare facility** – no. (%) | 7 (10.6%) | 16 (23.2%) | 23 (17%) | 0.08 |
| **Visit to healthcare facility within past week** – no. (%) | 13 (19.7%) | 22 (31.9%) | 35 (25.9%) | 0.18 |
| **Adherence to mask wearing** – no. (%) |  |  |  |  |
| - Always | 40 (60.6%) | 50 (72.5%) | 90 (66.7%) | 0.88 |
| - Often | 13 (19.7%) | 14 (20.3%) | 27 (20%) |  |
| - Sometimes | 2 (3%) | 1 (1.4%) | 3 (2.2%) |  |
| - Rarely | 0 (0%) | 0 (0%) | 0 (0%) |  |
| - Never | 1 (1.5%) | 1 (1.4%) | 2 (1.5%) |  |
| - Not reported | 10 (15.2%) | 3 (4.5%) | 13 (9.6%) |  |
| **Adherence to social distancing** – no. (%) |  |  |  |  |
| - Always | 28 (42.4%) | 46 (66.7%) | 74 (54.8%) | 0.03 |
| - Often | 27 (40.9%) | 17 (24.6%) | 44 (32.6%) |  |
| - Sometimes | 1 (1.5%) | 3 (4.3%) | 4 (3%) |  |
| - Rarely | 0 (0%) | 0 (0%) | 0 (0%) |  |
| - Never | 0 (0%) | 0 (0%) | 0 (0%) |  |
| - Not reported | 10 (15.2%) | 3 (4.5%) | 13 (9.6%) |  |
| **Adherence to handwashing** – no. (%) | | | | |
| - Always | 31 (47%) | 50 (72.5%) | 81 (60%) | 0.01 |
| - Often | 21 (31.8%) | 9 (13%) | 30 (22.2%) |  |
| - Sometimes | 4 (6.1%) | 5 (7.2%) | 9 (6.7%) |  |
| - Rarely | 0 (0%) | 2 (2.9%) | 2 (1.5%) |  |
| - Never | 0 (0%) | 0 (0%) | 0 (0%) |  |
| - Not reported | 10 (15.2%) | 3 (4.5%) | 13 (9.6%) |  |

**Table S2c.** COVID-19 exposure risk factors, mITTasymptomatic analysis.

|  | **LGG** | **Placebo** | **All Participants** |  |
| --- | --- | --- | --- | --- |
|  | **N=52**  **(50%)** | **N=52**  **(50%)** | **N=104**  **(100%)** | **P-Value** |
| **Household size** – no. (%) | | | | |
| - 2 | 9 (17.3%) | 20 (38.5%) | 29 (27.9%) | 0.41 |
| - 3 | 7 (13.5%) | 6 (11.5%) | 13 (12.5%) |  |
| - 4 | 11 (21.2%) | 11 (21.2%) | 22 (21.2%) |  |
| - 5 | 10 (19.2%) | 8 (15.4%) | 18 (17.3%) |  |
| - ≥6 | 10 (19.2) | 5 (9.6%) | 15 (14.4%) |  |
| - Not reported | 5 (9.6%) | 2 (3.8%) | 7 (6.7%) |  |
| **COVID-positive contacts in household** – no. (%) | | | | |
| - 1 | 24 (46.2%) | 19 (36.5%) | 43 (41.3%) | 0.44 |
| - 2 | 8 (15.4%) | 5 (9.6%) | 13 (12.5%) |  |
| - 3 | 1 (1.9%) | 1 (1.9%) | 2 (1.9%) |  |
| - 4 | 0 (0%) | 2 (3.8%) | 2 (1.9%) |  |
| - Not reported | 19 (36.5%) | 25 (48.1%) | 44 (42.3%) |  |
| **Close contact with COVID-positive person outside home within past week** – no (%) | 8 (15.4%) | 10 (19.2%) | 18 (17.3%) | 0.78 |
| **Participation in large social gatherings** **within past week** – no. (%) | 3 (5.8%) | 6 (11.5%) | 9 (8.7%) | 0.38 |
| **Employment outside of home** – no. (%) | 18 (34.6%) | 20 (38.5%) | 38 (36.5%) | 0.99 |
| **Employment at a healthcare facility** – no. (%) | 2 (3.8%) | 12 (23.1%) | 14 (13.5%) | 0.007 |
| **Visit to healthcare facility within past week** – no. (%) | 10 (19.2%) | 16 (30.8%) | 26 (25%) | 0.28 |
| **Adherence to mask wearing** – no. (%) |  |  |  |  |
| - Always | 32 (61.5%) | 37 (71.2%) | 69 (66.3%) | 0.92 |
| - Often | 9 (17.3%) | 10 (19.2%) | 19 (18.3%) |  |
| - Sometimes | 2 (3.8%) | 1 (1.9%) | 3 (2.9%) |  |
| - Rarely | 0 (0%) | 0 (0%) | 0 (0%) |  |
| - Never | 1 (1.9%) | 1 (1.9%) | 2 (1.9%) |  |
| - Not reported | 8 (15.4%) | 3 (5.8%) | 11 (10.6%) |  |
| **Adherence to social distancing** – no. (%) |  |  |  |  |
| - Always | 26 (50%) | 33 (63.5%) | 59 (56.7%) | 0.55 |
| - Often | 17 (32.7%) | 14 (26.9%) | 31 (29.8%) |  |
| - Sometimes | 1 (1.9%) | 2 (3.8%) | 3 (2.9%) |  |
| - Rarely | 0 (0%) | 0 (0%) | 0 (0%) |  |
| - Never | 0 (0%) | 0 (0%) | 0 (0%) |  |
| - Not reported | 8 (15.4%) | 3 (5.8%) | 11 (10.6%) |  |
| **Adherence to handwashing** – no. (%) |  |  |  |  |
| - Always | 25 (48.1%) | 35 (67.3%) | 60 (57.7%) | 0.10 |
| - Often | 16 (30.8%) | 8 (15.4%) | 24 (23.1%) |  |
| - Sometimes | 3 (5.8%) | 4 (7.7%) | 7 (6.7%) |  |
| - Rarely | 0 (0%) | 2 (3.8%) | 2 (1.9%) |  |
| - Never | 0 (0%) | 0 (0%) | 0 (0%) |  |
| - Not reported | 8 (15.4%) | 3 (5.8%) | 11 (10.6%) |  |

**Table S3a.** Self-reported symptoms, ITT analysis.

|  | **LGG** | **Placebo** | **All Participants** |  |
| --- | --- | --- | --- | --- |
|  | **N=91**  **(50%)** | **N=91**  **(50%)** | **N=182**  **(100%)** | **P-Value** |
| **Fever** | 2 (2.2%) | 4 (4.4%) | 6 (3.3%) | 0.46 |
| **Chills** | 3 (3.3%) | 6 (6.6%) | 9 (4.9%) | 0.36 |
| - Mild | 2 (2.2%) | 4 (4.4%) | 6 (3.3%) | >0.99 |
| - Moderate | 1 (1.1%) | 2 (2.2%) | 3 (1.6%) |  |
| - Severe | 0 (0%) | 0 (0%) | 0 (0%) |  |
| **Headache** | 10 (11%) | 17 (18.7%) | 27 (14.8%) | 0.20 |
| - Mild | 3 (3.3%) | 7 (7.7%) | 10 (5.5%) | 0.37 |
| - Moderate | 7 (7.7%) | 8 (8.8%) | 15 (8.2%) |  |
| - Severe | 0 (0%) | 2 (2.2%) | 2 (1.1%) |  |
| **Muscle aches** | 4 (4.4%) | 9 (9.9%) | 13 (7.1%) | 0.19 |
| - Mild | 2 (2.2%) | 7 (7.7%) | 9 (4.9%) | 0.32 |
| - Moderate | 2 (2.2%) | 2 (2.2%) | 4 (2.2%) |  |
| - Severe | 0 (0%) | 0 (0%) | 0 (0%) |  |
| **Runny nose** | 13 (14.3%) | 10 (11%) | 23 (12.6%) | 0.38 |
| - Mild | 10 (11%) | 5 (5.5%) | 15 (8.2%) | 0.18 |
| - Moderate | 3 (3.3%) | 5 (5.5%) | 8 (4.4%) |  |
| - Severe | 0 (0%) | 0 (0%) | 0 (0%) |  |
| **Sore throat** | 8 (8.8%) | 11 (12.1%) | 19 (10.4%) | 0.57 |
| - Mild | 7 (7.7%) | 7 (7.7%) | 14 (7.7%) | 0.24 |
| - Moderate | 1 (1.1%) | 4 (4.4%) | 5 (2.7%) |  |
| - Severe | 0 (0%) | 0 (0%) | 0 (0%) |  |
| **Cough** | 5 (5.5%) | 12 (13.2%) | 17 (9.3%) | 0.10 |
| - Mild | 3 (3.3%) | 8 (8.8%) | 11 (6%) | 0.43 |
| - Moderate | 2 (2.2%) | 2 (2.2%) | 4 (2.2%) |  |
| - Severe | 0 (0%) | 2 (2.2%) | 2 (1.1%) |  |
| **Shortness of breath** | 1 (1.1%) | 2 (2.2%) | 3 (1.6%) | 0.60 |
| - Mild | 0 (0%) | 2 (2.2%) | 2 (1.1%) | 0.08 |
| - Moderate | 1 (1.1%) | 0 (0%) | 1 (0.5%) |  |
| - Severe | 0 (0%) | 0 (0%) | 0 (0%) |  |
| **Nausea** | 2 (2.2%) | 5 (5.5%) | 7 (3.8%) | 0.29 |
| - Mild | 1 (1.1%) | 4 (4.4%) | 5 (2.7%) | 0.43 |
| - Moderate | 1 (1.1%) | 1 (1.1%) | 2 (1.1%) |  |
| - Severe | 0 (0%) | 0 (0%) | 0 (0%) |  |
| **Diarrhea** | 3 (3.3%) | 9 (9.9%) | 12 (6.6%) | 0.09 |
| - Mild | 1 (1.1%) | 2 (2.2%) | 3 (1.6%) | 0.36 |
| - Moderate | 2 (2.2%) | 3 (3.3%) | 5 (2.7%) |  |
| - Severe | 0 (0%) | 4 (4.4%) | 4 (2.2%) |  |
| **Bloating** | 8 (8.8%) | 14 (15.4%) | 22 (12.1%) | 0.23 |
| - Mild | 4 (4.4%) | 5 (5.5%) | 9 (4.9%) | 0.65 |
| - Moderate | 4 (4.4%) | 8 (8.8%) | 12 (6.6%) |  |
| - Severe | 0 (0%) | 1 (1.1%) | 1 (0.5%) |  |
| **Stomach upset or pain** | 8 (8.8%) | 15 (16.5%) | 23 (12.6%) | 0.16 |
| - Mild | 5 (5.5%) | 8 (8.8%) | 13 (7.1%) | 0.73 |
| - Moderate | 3 (3.3%) | 6 (6.6%) | 9 (4.9%) |  |
| - Severe | 0 (0%) | 1 (1.1%) | 1 (0.5%) |  |
| **Constipation** | 3 (3.3%) | 6 (6.6%) | 9 (4.9%) | 0.36 |
| - Mild | 3 (3.3%) | 2 (2.2%) | 5 (2.7%) | 0.06 |
| - Moderate | 0 (0%) | 4 (4.4%) | 4 (2.2%) |  |
| - Severe | 0 (0%) | 0 (0%) | 0 (0%) |  |
| **Loss of smell** | 2 (2.2%) | 8 (8.8%) | 10 (5.5%) | 0.06 |
| - Mild | 2 (2.2%) | 3 (3.3%) | 5 (2.7%) | 0.29 |
| - Moderate | 0 (0%) | 3 (3.3%) | 3 (1.6%) |  |
| - Severe | 0 (0%) | 2 (2.2%) | 2 (1.1%) |  |
| **Loss of taste** | 3 (3.3%) | 5 (5.5%) | 8 (4.4%) | 0.53 |
| - Mild | 3 (3.3%) | 0 (0%) | 3 (1.6%) | 0.02 |
| - Moderate | 0 (0%) | 3 (3.3%) | 3 (1.6%) |  |
| - Severe | 0 (0%) | 2 (2.2%) | 2 (1.1%) |  |
| **Other symptoms** | 2 (2.2%) | 2 (2.2%) | 4 (2.2%) | 0.94 |
| - Mild | 2 (2.2%) | 0 (0%) | 2 (1.1%) | 0.14 |
| - Moderate | 0 (0%) | 1 (1.1%) | 1 (0.5%) |  |
| - Severe | 0 (0%) | 1 (1.1%) | 1 (0.5%) |  |

**Table S3b.** Self-reported symptoms, mITTrt analysis.

|  | **LGG** | **Placebo** | **All Participants** |  |
| --- | --- | --- | --- | --- |
|  | **N=66**  **(48.9%)** | **N=69**  **(51.1%)** | **N=135**  **(100%)** | **P-Value** |
| **Fever** | 2 (3%) | 4 (5.8%) | 6 (4.4%) | 0.44 |
| **Chills** | 3 (4.5%) | 6 (8.7%) | 9 (6.7%) | 0.33 |
| - Mild | 2 (3%) | 4 (5.8%) | 6 (4.4%) | >0.99 |
| - Moderate | 1 (1.5%) | 2 (2.9%) | 3 (2.2%) |  |
| - Severe | 0 (0%) | 0 (0%) | 0 (0%) |  |
| **Headache** | 10 (15.2%) | 17 (24.6%) | 27 (20%) | 0.17 |
| - Mild | 3 (4.5%) | 7 (10.1%) | 10 (7.4%) | 0.37 |
| - Moderate | 7 (10.6%) | 8 (11.6%) | 15 (11.1%) |  |
| - Severe | 0 (0%) | 2 (2.9%) | 2 (1.5%) |  |
| **Muscle aches** | 4 (6.1%) | 9 (13%) | 13 (9.6%) | 0.17 |
| - Mild | 2 (3%) | 7 (10.1%) | 9 (6.7%) | 0.32 |
| - Moderate | 2 (3%) | 2 (2.9%) | 4 (3%) |  |
| - Severe | 0 (0%) | 0 (0%) | 0 (0%) |  |
| **Runny nose** | 13 (19.7%) | 10 (14.5%) | 23 (17%) | 0.42 |
| - Mild | 10 (15.2%) | 5 (7.2%) | 15 (11.1%) | 0.18 |
| - Moderate | 3 (4.5%) | 5 (7.2%) | 8 (5.9%) |  |
| - Severe | 0 (0%) | 0 (0%) | 0 (0%) |  |
| **Sore throat** | 8 (12.1%) | 11 (15.9%) | 19 (14.1%) | 0.52 |
| - Mild | 7 (10.6%) | 7 (10.1%) | 14 (10.4%) | 0.24 |
| - Moderate | 1 (1.5%) | 4 (5.8%) | 5 (3.7%) |  |
| - Severe | 0 (0%) | 0 (0%) | 0 (0%) |  |
| **Cough** | 5 (7.6%) | 12 (17.4%) | 17 (12.6%) | 0.09 |
| - Mild | 3 (4.5%) | 8 (11.6%) | 11 (8.1%) | 0.43 |
| - Moderate | 2 (3%) | 2 (2.9%) | 4 (3%) |  |
| - Severe | 0 (0%) | 2 (2.9%) | 2 (1.5%) |  |
| **Shortness of breath** | 1 (1.5%) | 2 (2.9%) | 3 (2.2%) | 0.59 |
| - Mild | 0 (0%) | 2 (2.9%) | 2 (1.5%) | 0.08 |
| - Moderate | 1 (1.5%) | 0 (0%) | 1 (0.7%) |  |
| - Severe | 0 (0%) | 0 (0%) | 0 (0%) |  |
| **Nausea** | 2 (3%) | 5 (7.2%) | 7 (5.2%) | 0.27 |
| - Mild | 1 (1.5%) | 4 (5.8%) | 5 (3.7%) | 0.43 |
| - Moderate | 1 (1.5%) | 1 (1.4%) | 2 (1.5%) |  |
| - Severe | 0 (0%) | 0 (0%) | 0 (0%) |  |
| **Diarrhea** | 3 (4.5%) | 9 (13%) | 12 (8.9%) | 0.08 |
| - Mild | 1 (1.5%) | 2 (2.9%) | 3 (2.2%) | 0.36 |
| - Moderate | 2 (3%) | 3 (4.3%) | 5 (3.7%) |  |
| - Severe | 0 (0%) | 4 (5.8%) | 4 (3%) |  |
| **Bloating** | 8 (12.1%) | 14 (20.3%) | 22 (16.3%) | 0.20 |
| - Mild | 4 (6.1%) | 5 (7.2%) | 9 (6.7%) | 0.65 |
| - Moderate | 4 (6.1%) | 8 (11.6%) | 12 (8.9%) |  |
| - Severe | 0 (0%) | 1 (1.4%) | 1 (0.7%) |  |
| **Stomach upset or pain** | 8 (12.1%) | 15 (21.7%) | 23 (17%) | 0.14 |
| - Mild | 5 (7.6%) | 8 (11.6%) | 13 (9.6%) | 0.73 |
| - Moderate | 3 (4.5%) | 6 (8.7%) | 9 (6.7%) |  |
| - Severe | 0 (0%) | 1 (1.4%) | 1 (0.7%) |  |
| **Constipation** | 3 (4.5%) | 6 (8.7%) | 9 (6.7%) | 0.33 |
| - Mild | 3 (4.5%) | 2 (2.9%) | 5 (3.7%) | 0.06 |
| - Moderate | 0 (0%) | 4 (5.8%) | 4 (3%) |  |
| - Severe | 0 (0%) | 0 (0%) | 0 (0%) |  |
| **Loss of smell** | 2 (3%) | 8 (11.6%) | 10 (7.4%) | 0.06 |
| - Mild | 2 (3%) | 3 (4.3%) | 5 (3.7%) | 0.27 |
| - Moderate | 0 (0%) | 3 (4.3%) | 3 (2.2%) |  |
| - Severe | 0 (0%) | 2 (2.9%) | 2 (1.5%) |  |
| **Loss of taste** | 3 (4.5%) | 5 (7.2%) | 8 (5.9%) | 0.51 |
| - Mild | 3 (4.5%) | 0 (0%) | 3 (2.2%) | 0.02 |
| - Moderate | 0 (0%) | 3 (4.3%) | 3 (2.2%) |  |
| - Severe | 0 (0%) | 2 (2.9%) | 2 (1.5%) |  |
| **Other symptoms** | 2 (3%) | 2 (2.9%) | 4 (3%) | 0.96 |
| - Mild | 2 (3%) | 0 (0%) | 2 (1.5%) | 0.14 |
| - Moderate | 0 (0%) | 1 (1.4%) | 1 (0.7%) |  |
| - Severe | 0 (0%) | 1 (1.4%) | 1 (0.7%) |  |

**Table S3c.** Self-reported symptoms, mITTasymptomatic analysis.

|  | **LGG** | **Placebo** | **All Participants** |  |
| --- | --- | --- | --- | --- |
|  | **N=52**  **(50%)** | **N=52**  **(50%)** | **N=104**  **(100%)** | **P-Value** |
| **Fever** | 0 (0%) | 2 (3.8%) | 2 (1.9%) | 0.15 |
| **Chills** | 1 (1.9%) | 3 (5.8%) | 4 (3.8%) | 0.31 |
| - Mild | 1 (1.9%) | 2 (3.8%) | 3 (2.9%) | 0.51 |
| - Moderate | 0 (0%) | 1 (1.9%) | 1 (1%) |  |
| - Severe | 0 (0%) | 0 (0%) | 0 (0%) |  |
| **Headache** | 6 (11.5%) | 11 (21.2%) | 17 (16.3%) | 0.18 |
| - Mild | 1 (1.9%) | 5 (9.6%) | 6 (5.8%) | 0.16 |
| - Moderate | 5 (9.6%) | 4 (7.7%) | 9 (8.7%) |  |
| - Severe | 0 (0%) | 2 (3.8%) | 2 (1.9%) |  |
| **Muscle aches** | 1 (1.9%) | 5 (9.6%) | 6 (5.8%) | 0.09 |
| - Mild | 1 (1.9%) | 5 (9.6%) | 6 (5.8%) | 0.09 |
| - Moderate | 0 (0%) | 0 (0%) | 0 (0%) |  |
| - Severe | 0 (0%) | 0 (0%) | 0 (0%) |  |
| **Runny nose** | 6 (11.5%) | 4 (7.7%) | 10 (9.6%) | 0.51 |
| - Mild | 3 (5.8%) | 3 (5.8%) | 6 (5.8%) | 0.43 |
| - Moderate | 3 (5.8%) | 1 (1.9%) | 4 (3.8%) |  |
| - Severe | 0 (0%) | 0 (0%) | 0 (0%) |  |
| **Sore throat** | 3 (5.8%) | 7 (13.5%) | 10 (9.6%) | 0.18 |
| - Mild | 3 (5.8%) | 5 (9.6%) | 8 (7.7%) | 0.30 |
| - Moderate | 0 (0%) | 2 (3.8%) | 2 (1.9%) |  |
| - Severe | 0 (0%) | 0 (0%) | 0 (0%) |  |
| **Cough** | 3 (5.8%) | 6 (11.5%) | 9 (8.7%) | 0.30 |
| - Mild | 2 (3.8%) | 4 (7.7%) | 6 (5.8%) | 0.69 |
| - Moderate | 1 (1.9%) | 1 (1.9%) | 2 (1.9%) |  |
| - Severe | 0 (0%) | 1 (1.9%) | 1 (1%) |  |
| **Shortness of breath** | 1 (1.9%) | 2 (3.8%) | 3 (2.9%) | 0.56 |
| - Mild | 0 (0%) | 2 (3.8%) | 2 (1.9%) | 0.08 |
| - Moderate | 1 (1.9%) | 0 (0%) | 1 (1%) |  |
| - Severe | 0 (0%) | 0 (0%) | 0 (0%) |  |
| **Nausea** | 1 (1.9%) | 4 (7.7%) | 5 (4.8%) | 0.17 |
| - Mild | 0 (0%) | 3 (5.8%) | 3 (2.9%) | 0.17 |
| - Moderate | 1 (1.9%) | 1 (1.9%) | 2 (1.9%) |  |
| - Severe | 0 (0%) | 0 (0%) | 0 (0%) |  |
| **Diarrhea** | 2 (3.8%) | 3 (5.8%) | 5 (4.8%) | 0.65 |
| - Mild | 1 (1.9%) | 0 (0%) | 1 (1%) | 0.33 |
| - Moderate | 1 (1.9%) | 2 (3.8%) | 3 (2.9%) |  |
| - Severe | 0 (0%) | 1 (1.9%) | 1 (1%) |  |
| **Bloating** | 5 (9.6%) | 8 (15.4%) | 13 (12.5%) | 0.37 |
| - Mild | 3 (5.8%) | 2 (3.8%) | 5 (4.8%) | 0.39 |
| - Moderate | 2 (3.8%) | 5 (9.6%) | 7 (6.7%) |  |
| - Severe | 0 (0%) | 1 (1.9%) | 1 (1%) |  |
| **Stomach upset or pain** | 5 (9.6%) | 9 (17.3%) | 14 (13.5%) | 0.25 |
| - Mild | 3 (5.8%) | 4 (7.7%) | 7 (6.7%) | 0.58 |
| - Moderate | 2 (3.8%) | 5 (9.6%) | 7 (6.7%) |  |
| - Severe | 0 (0%) | 0 (0%) | 0 (0%) |  |
| **Constipation** | 2 (3.8%) | 4 (7.7%) | 6 (5.8%) | 0.40 |
| - Mild | 2 (3.8%) | 2 (3.8%) | 4 (3.8%) | 0.22 |
| - Moderate | 0 (0%) | 2 (3.8%) | 2 (1.9%) |  |
| - Severe | 0 (0%) | 0 (0%) | 0 (0%) |  |
| **Loss of smell** | 1 (1.9%) | 6 (11.5%) | 7 (6.7%) | 0.05 |
| - Mild | 1 (1.9%) | 1 (1.9%) | 2 (1.9%) | 0.23 |
| - Moderate | 0 (0%) | 3 (5.8%) | 3 (2.9%) |  |
| - Severe | 0 (0%) | 2 (3.8%) | 2 (1.9%) |  |
| **Loss of taste** | 2 (3.8%) | 5 (9.6%) | 7 (6.7%) | 0.24 |
| - Mild | 2 (3.8%) | 0 (0%) | 2 (1.9%) | 0.03 |
| - Moderate | 0 (0%) | 3 (5.8%) | 3 (2.9%) |  |
| - Severe | 0 (0%) | 2 (3.8%) | 2 (1.9%) |  |
| **Other symptoms** | 1 (1.9%) | 2 (3.8%) | 3 (2.9%) | 0.56 |
| - Mild | 1 (1.9%) | 0 (0%) | 1 (1%) | 0.22 |
| - Moderate | 0 (0%) | 1 (1.9%) | 1 (1%) |  |
| - Severe | 0 (0%) | 1 (1.9%) | 1 (1%) |  |

**Table S4.** Sensitivity analysis by sex, ITT, mITTrt, and mITTasymptomatic analyses.

|  | **Female** | **Male** | **All Participants** | **P-Value** |
| --- | --- | --- | --- | --- |
| **ITT** | **N=115 (63.2%)** | **N=67 (36.8%)** | **N = 182 (100%)** |  |
| **Any symptoms** – no. (%) | 41 (35.7%) | 22 (32.8%) | 63 (34.6%) | 0.70 |
| **Any moderate/severe symptoms** – no. (%) | 29 (25.2%) | 11 (16.4%) | 40 (22%) | 0.17 |
| **Reported COVID-19 Diagnosis** – no. (%) | 12 (10.4%) | 10 (14.9%) | 22 (12.1%) | 0.37 |
| **mITTrt** | **N=87 (64.4%)** | **N=48(35.6%)** | **N = 135 (100%)** |  |
| **Any symptoms** – no. (%) | 41 (47.1%) | 22 (45.8%) | 63 (46.7%) | 0.88 |
| **Any moderate/severe symptoms** – no. (%) | 29 (33.3%) | 11 (22.9%) | 40 (29.6%) | 0.20 |
| **Reported COVID-19 Diagnosis** – no. (%) | 10 (11.5%) | 9 (18.8%) | 19 (14.1%) | 0.25 |
| **mITTasymptomatic** | **N=63**  **(60.6%)** | **N=41 (39.4%)** | **N=104**  **(100%)** |  |
| **Any symptoms** – no. (%) | 23 (36.5%) | 16 (39%) | 39 (37.5%) | 0.80 |
| **Any moderate/severe symptoms** – no. (%) | 16 (25.4%) | 9 (22%) | 25 (24%) | 0.69 |
| **Reported COVID-19 Diagnosis** – no. (%) | 8 (8.8%) | 14 (15.4%) | 22 (12.1%) | 0.17 |

**Table S5.** Sensitivity analysis by age, ITT/mITTrt/mITTasymptomatic analysis.

|  | **Age <18** | **Age 18-64** | **Age >=65** | **All Participants** | **P-Value** |
| --- | --- | --- | --- | --- | --- |
| **ITT** | **N=41**  **(22/5%)** | **N=131**  **(72.0%)** | **N=10**  **(5.5%)** | **N=182**  **(100%)** |  |
| **Any symptoms** – no. (%) | 6 (14.6%) | 51 (38.9%) | 6 (60%) | 63 (34.6%) | 0.004 |
| **Any moderate/severe symptoms** – no. (%) | 1 (2.4%) | 37 (28.2%) | 2 (20%) | 40 (22%) | 0.002 |
| **Reported COVID-19 Diagnosis** – no. (%) | 5 (12.2%) | 13 (9.9%) | 4 (40%) | 22 (12.1%) | 0.02 |
| **mITTrt** | **N=27**  **(20%)** | **N=99**  **(73.3%)** | **N=9**  **(6.7%)** | **N=135**  **(100%)** |  |
| **Any symptoms** – no. (%) | 6 (22.2%) | 51 (51.5%) | 6 (66.7%) | 63 (46.7%) | 0.02 |
| **Any moderate/severe symptoms** – no. (%) | 1 (3.7%) | 37 (37.4%) | 2 (22.2%) | 40 (29.6%) | 0.003 |
| **Reported COVID-19 Diagnosis** – no. (%) | 2 (7.4%) | 13 (13.1%) | 4 (44.4%) | 19 (14.1%) | 0.02 |
| **mITTasymptomatic** | **N=25**  **(24.0%)** | **N=72**  **(69.2%)** | **N=7**  **(6.7%)** | **N=104**  **(100%)** |  |
| **Any symptoms** – no. (%) | 4 (16%) | 31 (43.1%) | 4 (57.1%) | 39 (37.5%) | 0.03 |
| **Any moderate/severe symptoms** – no. (%) | 1 (4%) | 23 (31.9%) | 1 (14.3%) | 25 (24%) | 0.02 |
| **Reported COVID-19 Diagnosis** – no. (%) | 1 (4%) | 7 (9.7%) | 3 (42.9%) | 11 (10.6%) | 0.01 |

**Table S6.** Logistic regression model for any symptoms at D28.

| **Variable** | **Odds Ratio** | **95% Confidence Interval** | **P-Value** | **Overall P-Value** |
| --- | --- | --- | --- | --- |
|  | Randomization Arm | | | |
| Placebo | -REF- |  |  | 0.06 |
| Active | 0.49 | 0.23 - 1.02 |  |  |
|  | Age group | | | |
| 18-64 | -REF- |  |  | 0.04 |
| <18 | 0.31 | 0.11 - 0.85 | 0.02 |  |
| >=65 | 1.61 | 0.37 - 6.99 | 0.52 |  |
|  | Current smoker | | | |
| No | -REF- |  |  | 0.98 |
| Yes | 0.99 | 0.33 - 2.98 |  |  |

**Table S7a/b/c.** Participant-Reported Adherence and Adverse Effects.

a. **ITT analysis.**

|  | **LGG** | **Placebo** | **All Participants** |  |
| --- | --- | --- | --- | --- |
|  | **N=91**  **(50%)** | **N=91**  **(50%)** | **N=182**  **(100%)** | **P-Value** |
| **Participant adherence outcomes** – no. (%) |  |  |  |  |
| - Did not report receiving study product | 25 (27.5%) | 22 (24.2%) | 47 (25.8%) | 0.83 |
| - Received product; did not report adherence | 13 (14.3%) | 12 (13.2%) | 25 (13.7%) |  |
| - Received product and reported adherence | 53 (58.2%) | 57 (62.6%) | 110 (60.4%) |  |
| **Median percent adherence*** – % (IQR) | 100 (93-100) | 100 (93-100) | 100 (93-100) | 0.82 |
| **Perceived study product assignment - no. (%)** |  |  |  |  |
| - LGG | 15 (16.5%) | 11 (12.1%) | 26 (14.3%) | 0.29 |
| - Placebo | 5 (5.5%) | 10 (11%) | 15 (8.2%) |  |
| - Unknown | 29 (31.9%) | 25 (27.5%) | 54 (29.7%) |  |
| **Attributed symptoms to study product** – no. (%) | 8 (8.8%) | 12 (23.1%) | 20 (19.2%) | 0.35 |
| - Nausea | 0 (0%) | 0 (0%) | 0 (0%) | -- |
| - Bloating | 7 (7.7%) | 8 (8.8%) | 15 (8.2%) | 0.79 |
| - Stomach Upset | 2 (2.2%) | 2 (2.2%) | 4 (2.2%) | 1.00 |
| - Diarrhea | 1 (1.1%) | 2 (2.2%) | 3 (1.6%) | 0.56 |
| - Constipation | 1 (1.1%) | 3 (3.3%) | 4 (2.2%) | 0.31 |
| **Stopped study product due to symptoms** – no. (%) | 0 (0%) | 5 (5.5%) | 5 (2.7%) | 0.02 |

* Median adherence was calculated for participants who reported at least one adherence time point.

b. **mITTrt analysis.**

|  | **LGG** | **Placebo** | **All Participants** |  |
| --- | --- | --- | --- | --- |
|  | **N=66**  **(48.9%)** | **N=69**  **(51.1%)** | **N=135**  **(100%)** | **P-Value** |
| Median percent adherence* - % (IQR) | 100 (93 - 100) | 100 (93 - 100) | 100 (93 - 100) | 0.82 |
| **Perceived study product assignment - no. (%)** | | | | |
| - LGG | 15 (22.7%) | 11 (15.9%) | 26 (19.3%) | 0.29 |
| - Placebo | 5 (7.6%) | 10 (14.5%) | 15 (11.1%) |  |
| - Unknown | 29 (43.9%) | 25 (36.2%) | 54 (40%) |  |
| **Attributed symptoms to study product** – no. (%) | 8 (12.1%) | 12 (17.4%) | 20 (14.8%) | 0.39 |
| - Nausea | 0 (0%) | 0 (0%) | 0 (0%) | -- |
| - Bloating | 7 (10.6%) | 8 (11.6%) | 15 (11.1%) | 0.86 |
| - Stomach Upset | 2 (3.0%) | 2 (2.9%) | 4 (3.0%) | 1.00 |
| - Diarrhea | 1 (1.5%) | 2 (2.9%) | 3 (2.2%) | 0.59 |
| - Constipation | 1 (1.5%) | 3 (4.3%) | 4 (3.0%) | 0.34 |
| **Stopped study product due to symptoms** – no. (%) | 0 (0%) | 5 (7.2%) | 5 (4.8%) | 0.03 |

*Median adherence was calculated for participants who reported at least one adherence time point.

c. **mITTasymptomatic analysis.**

|  | **LGG** | **Placebo** | **All Participants** |  |
| --- | --- | --- | --- | --- |
|  | **N=52**  **(50%)** | **N=52**  **(50%)** | **N=104**  **(100%)** | **P-Value** |
| Median percent adherence* - % (IQR) | 100 (93 - 100) | 100 (96 - 100) | 100 (93 - 100) | 0.85 |
| **Perceived study product assignment - no. (%)** | | | | |
| - LGG | 9 (17.3%) | 9 (17.3%) | 18 (17.3%) | 0.67 |
| - Placebo | 5 (9.6%) | 7 (13.5%) | 12 (11.5%) |  |
| - Unknown | 24 (46.2%) | 19 (36.5%) | 43 (41.3%) |  |
| **Attributed symptoms to study product** – no. (%) | 8 (15.3%) | 12 (23.1%) | 20 (19.2%) | 0.32 |
| - Nausea | 0 (0%) | 0 (0%) | 0 (0%) | -- |
| - Bloating | 7 (13.4%) | 8 (15.3%) | 15 (14.4%) | 0.78 |
| - Stomach Upset | 2 (3.8%) | 2 (3.8%) | 4 (3.8%) | 1.0 |
| - Diarrhea | 1 (1.9 %) | 2 (3.8%) | 3 (2.9%) | 0.56 |
| - Constipation | 1 (1.9 %) | 3 (5.8%) | 4 (3.8%) | 0.31 |
| **Stopped study product due to symptoms** – no. (%) | 0 (0%) | 5 (9.6%) | 5 (4.8%) | 0.02 |

*Median adherence was calculated for participants who reported at least one adherence time point.
